# Racial and ethnic and sex differences in at-home estimates of sleep-disordered breathing parameters among Mexican American, Black, and Non-Hispanic White adults

**DOI:** 10.1101/2024.10.23.24315996

**Authors:** Yue Leng, Clémence Cavaillès, Carrie Peltz, Sid E. O’Bryant, Susan Redline, Kristine Yaffe

**Affiliations:** Department of Psychiatry and Behavioral Sciences, University of California, San Francisco, CA, USA; San Francisco Veterans Affairs Health Care System, San Francisco, CA, USA; University of North Texas Health Science Center, Fort Worth, USA; Division of Sleep and Circadian Disorders, Department of Medicine, Brigham and Women’s Hospital, Boston, MA, USA; Division of Sleep Medicine, Harvard Medical School, Boston, MA, USA; Departments of Neurology, Epidemiology and Biostatistics, University of California, San Francisco, CA, USA

**Keywords:** Sleep-disordered breathing, REM sleep, NREM sleep, race/ethnicity, disparities

## Abstract

**Background**

Racial and ethnic and sex differences in sleep may exist, but there are limited data directly comparing objective estimates of sleep-disordered breathing (SDB), particularly in rapid eye movement (REM) versus non-rapid eye movement (NREM) sleep, among Black, Mexican American (MA) and non-Hispanic White (NHW) men and women. Our goal is to investigate health disparities in SDB in a new, diverse cohort of older adults.

**Research Question**

Do SDB parameters during REM and NREM sleep differ by race and ethnicity or sex in community-dwelling older adults?.

**Methods**

The Dormir Study conducted a comprehensive sleep examination among eligible participants enrolled in the ongoing community-based Health and Aging Brain Study-Health Disparities (HABS-HD) cohort (2020-4), among Black, MA, and NHW adults aged 50 years and older. Here we characterize racial and ethnic and sex differences in SDB indices assessed by an FDA-approved Peripheral Arterial Tonometry (PAT)-based home sleep testing system.

**Results**

We examined 821 participants, including 543 (66.1%) women, and 284 (34.6%) MA and 174 (21.2%) Black individuals, with a mean age of 66.6±8.5 years. Around half (50.5%) of the participants had moderate to severe SDB as defined by the respiratory event index (REI based on 3% desaturations) of ≥15/ hour, 72.7% with moderate to severe REM SDB (REM-REI ≥ 15/hour) and 39.5% with moderate to severe NREM SDB (NREM-REI ≥15/hour). The prevalence of SDB did not differ by race or sex. However, significant racial and ethnic and sex differences were observed for REM-specific SDB metrics. Overall, Black women had the highest REM REI, and NHW men had the lowest REM REI and REM ODI. After controlling for age, sex, education, income, employment status, cognitive status, BMI, history of hypertension, diabetes, stroke, and coronary heart disease, and sleep medication use, Black participants had a REM-REI that was 3 events per hour higher than that of NHW adults, while NREM-REI were similar. MA individuals had similar REM or NREM SDB parameters compared to NHW adults but exhibited higher average blood oxygen levels.

**Conclusions**

In this community-based cohort of middle- to older-aged NHW, MA, and Black adults, PAT-based measures of in-home sleep indicate a higher prevalence of REM SDB in Black adults, particularly Black women, compared to their NHW counterparts; this contrasts with the similar NREM parameters observed across racial and ethnic groups. Given the close link between REM SDB and adverse health outcomes, clinicians should pay more attention to this sleep apnea phenotype, especially in minoritized populations.

## INTRODUCTION

Sleep-disordered breathing (SDB), a common sleep disorder associated with numerous adverse health outcomes, affects nearly 1 billion adults aged 30-69 years globally^1–3^. Despite its widespread impact, most SDB cases remain undiagnosed and untreated, particularly in resource-limited settings, leading to significant health consequences^1,4^. Disparities in sleep health have recently emerged as a critical public health issue, as it is hypothesized that racial/ethnic minorities, such as Black and Mexican American (MA) individuals may experience worse sleep health due to higher comorbidities and lower socio-economic status (SES)^5^. However, disparities in SDB are poorly understood. Most studies rely on subjective metrics, which are inadequate for characterizing SDB. Although some research has objectively measured SDB, these studies are predominantly in older populations, with few including MA participants^6–8^.

No study has comprehensively compared SDB in rapid eye movement (REM) compared to non-REM (NREM) sleep among Black, MA, and non-Hispanic White (NHW) adults. REM sleep is often associated with more severe SDB, characterized by longer obstructive events with deeper desaturations ^9^. Several studies suggest that the greater “toxicity” of REM SDB events explain the observed strong association between REM SDB with adverse cardiovascular, cognitive, and metabolic outcomes^10^. While REM-predominant SDB has been reported to be more common in females compared to males, fewer data have examined race and ethnic differences in REM SDB. Notably, mechanisms underlying the pathogenesis of SDB (“endotypes”) appear to vary both by sex as well as by race and ethnicity. Specifically, women and older Black adults have been reported to have decreased pharyngeal collapsibility during sleep-which may provide relative protection from airway obstruction during NREM but not in REM sleep, when cholinergic-mediated inhibition of upper airway muscle dilators and reduced hypoxic and hypercapnic ventilatory responses may drive airway obstruction ^11^. Despite these findings, there remains a lack of research to characterize and compare SDB during REM and NREM sleep among men and women in community-settings^12^; few studies have examined the joint effects of race and sex on SDB.

Importantly, disparities in sleep health may contribute to broader health disparities, including an increased risk of cardiometabolic diseases and cognitive impairment^13–15^. Given that SDB events during REM sleep are often longer, more frequent, and are associated with greater oxyhemoglobin desaturation and higher cardiovascular disease risk compared to SDB during NREM sleep^16^, characterizing SDB in REM and NREM sleep among Black and MA men and women in a home setting has substantial public health implications^17^.

We therefore aimed to comprehensively characterize in-home SDB among community-dwelling middle- to older-aged MA, Black, and NHW men and women, with a focus on potential racial and ethnic and sex differences. We also examined if disparities in SDB among diverse populations are independent of SES and comorbidities.

## METHODS

The Health and Aging Brain Study-Health Disparities (HABS-HD) ^18^ is an ongoing, longitudinal community-based multi-ethnic study of brain aging among MA, Black, and NHW participants, who were recruited using a community-based participatory research (CBPR) approach and completed the baseline study visit. Participants need to be fluent in English or Spanish, willing to provide blood samples, and be capable of undergoing neuroimaging studies. Detailed inclusion and exclusion criteria have been described previously ^18^.

The Dormir Study, an ancillary HABS-HD study, was conducted between 2020 and 2024 and all eligible HABS-HD participants aged 50 years and older were invited for a comprehensive sleep examination, including single overnight assessment using a Type III home sleep apnea test (WatchPAT) and 7-day actigraphy and sleep diary completion. Participants who consented for re-contact from the ongoing HABS-HD cohort were first invited for a brief pre-screening questionnaire if interested. Those with permanent pacemakers (atrial pacing or VVI without sinus rhythm) and those currently using continuous positive airway pressure machine were excluded. Here, we focus on 821 participants with complete demographical and WatchPAT sleep data as of June 2024. All study participants provided written informed consent, and the Dormir Study was approved by the University of North Texas Health Science Center Institutional Review Board.

### At home sleep assessment using WatchPAT

During the at home assessment, participants used the WatchPAT 200 (Itamar Medical Ltd., Caesarea, Israel) for one night. Studies of less than 4 hours in duration were repeated. The WatchPAT 200 (Zoll Itamar Medical, IS) is an FDA-approved wrist-worn device that utilizes a plethysmographic based finger-mounted probe to record the PAT™ (Peripheral Arterial Tone) signal, heart rate, and oxygen saturation, and a built-in actigraph to measure movement and estimate sleep time. The PAT™ signal ^19^ measures pulsatile volume changes in the fingertip arterioles which reflects the relative state of the arterial vasomotor activity, and thus indirectly the level of sympathetic activation. When respiratory events conclude, a surge in sympathetic nervous system activity occurs, marked by an increased heart rate and decreased oxygen levels. The body position sensor uses a 3-axis accelerometer to indicate the patient’s sleeping posture (supine, prone, left, right, and sit). An integrated snore sensor uses an acoustic decibel detector with a highly sensitive microphone that responds to snoring and other audio range sounds, converting them into a small analog voltage, thereby providing measures of snoring sounds. The WatchPAT software detects apnea and hypopnea events by a proprietary algorithm based on identification of changes in peripheral tonometry, movement, heart rate, desaturation and snoring. Additionally, the software provides indices of overnight oxygen saturation, and using the heart rate, movement and arterial tonometry signals, distinguishes light from REM sleep with good accuracy^20^.

To generate the sleep exposure data for this study, the WatchPAT data were electronically transmitted to a Sleep Reading Center (Brigham and Women’s Hospital, Boston, MA) where the overnight records were scored by a trained technician who was blinded to other data. Review and scoring included quality assessment of each channel, identification of urgent alerts, and manual editing of respiratory events. Itamar’s zzzPAT software (versions 5.2.79.7p and 4.6.71.7, according to the calendar dates of the study) was first used for automatic analysis of respiratory events and sleep. Records were then manually annotated to ensure that all respiratory events and sleep stages reflected criteria proposed by Zhang et al^21^. In brief, events in estimated NREM sleep were retained if they showed a heart rate increase, a PAT decrease, and 3 ≥ percent desaturation, with no postural change. In estimated REM sleep, events were retained when associated with a ≥ 4% desaturation. Compared to polysomnography, this approach was ≥ reported to improve estimation of the Respiratory Event Index (REI; the number of breathing pauses per hour of estimated sleep; r=0.81), total sleep time (r=0.73) and REM time (r=0.64)^21^. SDB parameters of interest include REI, REM-REI, NREM-REI, oxygen desaturation index (ODI, the average number of ≥ 4% desaturation episodes per hour), REM-ODI, NREM-ODI, lowest ≥ oxygen saturation (SpO2), the percentage of sleep time with SpO2<90%, and the percentage of sleep time with snoring volume >40dB.

### Cognitive diagnoses and other measurements

Cognitive diagnoses, including normal control, mild cognitive impairment (MCI), and dementia, were determined using an algorithmic approach (decision tree) and subsequently confirmed through a consensus review^18^. Self-reported age, sex, education, income, employment status, sleep medication use, and medical history, including hypertension, diabetes, stroke, and coronary heart disease (CHD), were collected from clinical interviews. Body mass index (BMI) was calculated based on weight and height (kg/m2) assessed at the clinical interview.

### Statistical analysis

Participants’ demographic data, comorbidities, and sleep-related characteristics were described and compared by race and ethnicity using chi-squared tests for categorical variables, ANOVA tests for normally distributed continuous variables, and Kruskal-Wallis tests for skewed continuous variables. SDB characteristics were also compared by sex using t-tests for normally distributed continuous variables, and Mann-Whitney U tests for skewed continuous variables. Additionally, comparisons by race after stratifying by sex (six groups) were conducted using the same statistical tests as the initial race and ethnicity comparisons.

We examined the multivariable association between sleep characteristics and race and ethnicity using linear regression models adjusted for age, sex, education, income, employment status, cognitive status, BMI, history of hypertension, diabetes, stroke, and CHD, and sleep medication use. Additional adjustment for sleep time spent in supine position was also made to assess the influence of body position on SDB parameters. Cube and log transformations were applied for non-normally distributed variables, and results were back-transformed to the original scale. Results are displayed as adjusted means with their 95% confidence intervals. All statistical tests were two-sided, and analyses were performed using R version 4.4.0.

## RESULTS

Of the 821 Dormir participants, the mean age was 66.6±8.5 years, 543 (66.1%) were women, 176 (21.4%) had MCI or dementia, and 284 (34.6%) were MA and 174 (21.2%) Black individuals. Black or MA participants were younger, more likely to be women, had higher BMI, and were more likely to have MCI or dementia, or have a history of hypertension or diabetes (Supplemental Table 1). Table 1 shows the SDB characteristics by race and ethnicity. On average, the prevalence of SDB (REI≥ 5) was 87.8%, and about half (50.5%) of the sample had ≥ moderate to severe SDB (REI≥ 15). The proportion of participants with moderate to severe REM SDB (REM REI ≥ 15) and moderate to severe NREM SDB (NREM REI≥ 15) was 72.7% and 39.5%, respectively. While the proportions of moderate to severe SDB across REM and NREM were similar among Black (54.0%), MA (51.4%), and NHW (50.5%) participants, REM-specific indices of SDB showed that Black or MA participants had significantly higher REM REI and higher REM ODI. Besides, MA participants had higher average SpO2 levels, and Black and MA individuals had a greater percentage of time spent with snoring volume >40dB (p<0.003 for all above metrics).

**Table 1.** Description of sleep-disordered breathing parameters according to race and ethnicity.

| **Sleep-disordered breathing variables** | Total sample<br>(n=821) | Non-Hispanic<br>White<br>(n=363) | Mexican<br>American<br>(n=284) | Black<br>(n=174) | p-value<sup>a</sup> |
| --- | --- | --- | --- | --- | --- |
| REI | 15.0 (8.5,25.1) | 14.5 (7.9,24.6) | 15.6 (8.8,25.7) | 16.3 (9.6,24.4) | 0.20 |
| NREM-REI | 11.7 (5.9,20.8) | 11.1 (5.8,20.9) | 12.3 (5.9,21.7) | 12.0 (6.1,20.0) | 0.76 |
| REM-REI | 25.1 (13.6,37.45) | 22.7 (11.9,34.7) | 26.8 (15.0,38.3) | 27.7 (18.2,41.7) | 0.0006 |
| ODI | 7.1 (3.1,14.5) | 6.2 (2.8,13.1) | 8.1 (3.7,16.2) | 7.1 (3.5,13.6) | 0.04 |
| NREM-ODI | 4.6 (1.9,10.6) | 4.2 (1.7,10.1) | 5.5 (2.3,11.5) | 4.2 (1.9,9.1) | 0.10 |
| REM-ODI | 13.3 (5.7,25.85) | 11.4 (4.3,22.7) | 15.9 (6.5,28.8) | 14.3 (7.2,26.6) | 0.003 |
| Average SpO<sub>2</sub>, %* | 93.7 (±1.6) | 93.5 (±1.6) | 94.2 (±1.6) | 93.2 (±1.6) | <.0001 |
| SpO<sub>2</sub><90%, % | 0.3 (0.0,1.6) | 0.3 (0.0,1.6) | 0.2 (0.0,1.3) | 0.4 (0.0,2.4) | 0.13 |
| Snoring volume>40 dB, % | 13.4 (6.1,28.3) | 10.9 (5.7,24.1) | 15.1 (6.9,31.6) | 17.0 (7.6,35.0) | 0.0009 |

mean (± standard deviation); all other results are shown as n (%) or median (Q1,Q3)
Events in estimated NREM sleep were retained if they showed a heart rate increase, a PAT decrease, and ≥ 3 percent desaturation, with no postural change. In estimated REM sleep, events were retained when associated with a ≥ 4% desaturation.
<sup>a</sup> Mann-Whitney U test was used for continuous variables with a normal distribution and ANOVA test for continuous variables without a normal distribution, Chi-square test for categorical variables

The prevalence of SDB, not specified by sleep state and including moderate to severe SDB, did not differ by sex (Supplemental Table 2). Figure 1 and Supplemental Table 3 summarize the distribution of SDB characteristics by race and ethnicity and sex. Of the six groups, Black women had the highest REM REI and the highest proportion of snoring volume>40 dB; MA women had the highest average SpO2 levels; and NHW men had the lowest REM REI and REM ODI.

**Figure 1.**
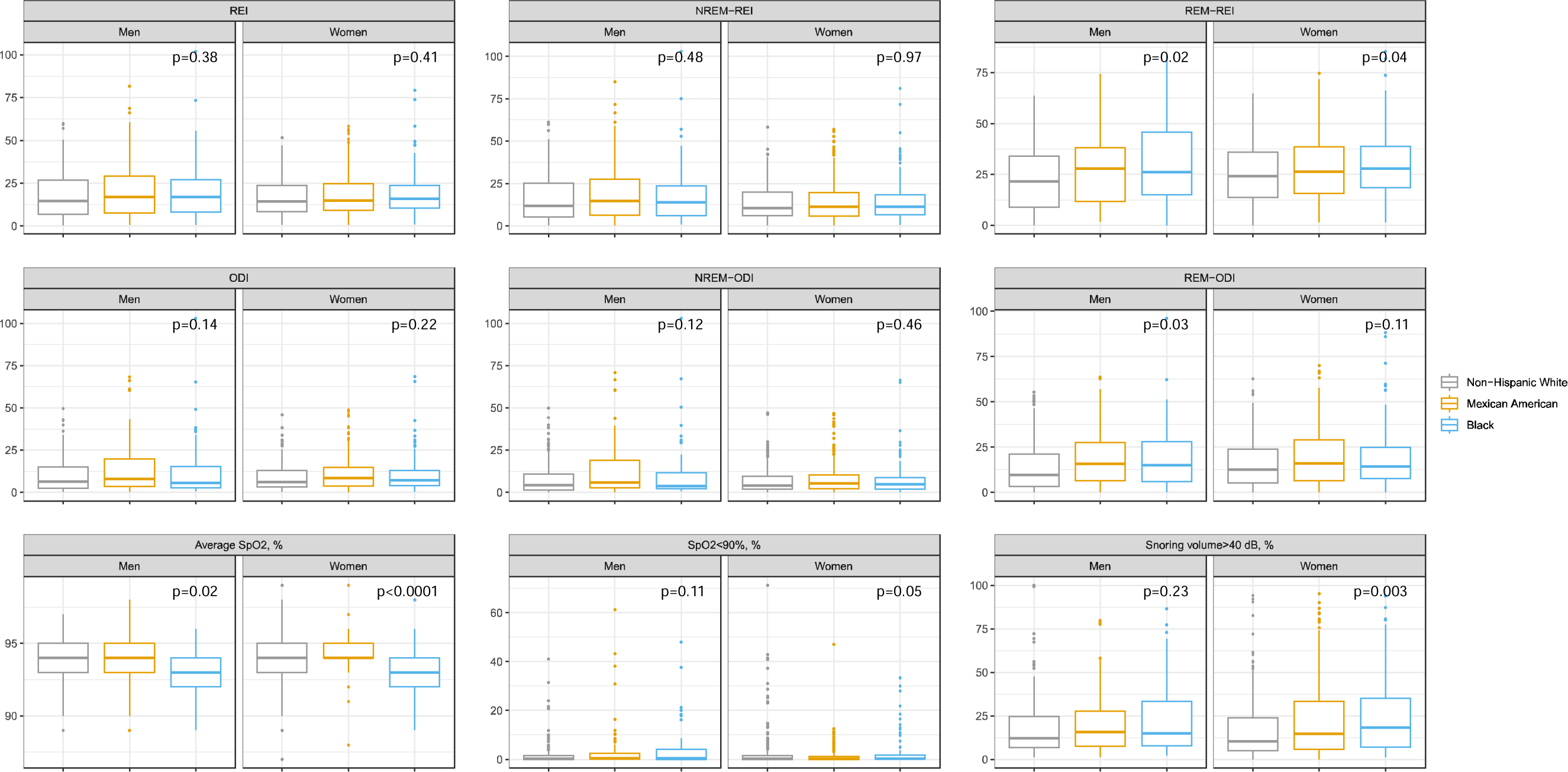
Sleep-disordered breathing parameters by race and sex. REI: Respiratory Event Index; REM: Rapid eye movement; NREM: Non-rapid eye movement; ODI: Oxygen desaturation index.

Figure 2 and Supplemental Table 4 show the adjusted mean sleep characteristics in each race and ethnicity group, after controlling for age, sex, education, income, employment status, cognitive status, BMI, history of hypertension, diabetes, stroke, and CHD, and sleep medication use. Compared to NHW participants, Black participants had significantly higher REM-REI, and MA participants had higher average SpO2 levels and lower percentage of sleep time spent with SpO2<90%. The adjusted mean REM-REI values were 23.4, 20.8, and 20.4 for Black, MA, and NHW individuals, respectively. Additional adjustment for sleep time spent in supine position did not appreciably alter the results. Other SDB characteristics, including overall REI, ODI, or snoring volume did not differ among the three race and ethnicity groups.

**Figure 2.**
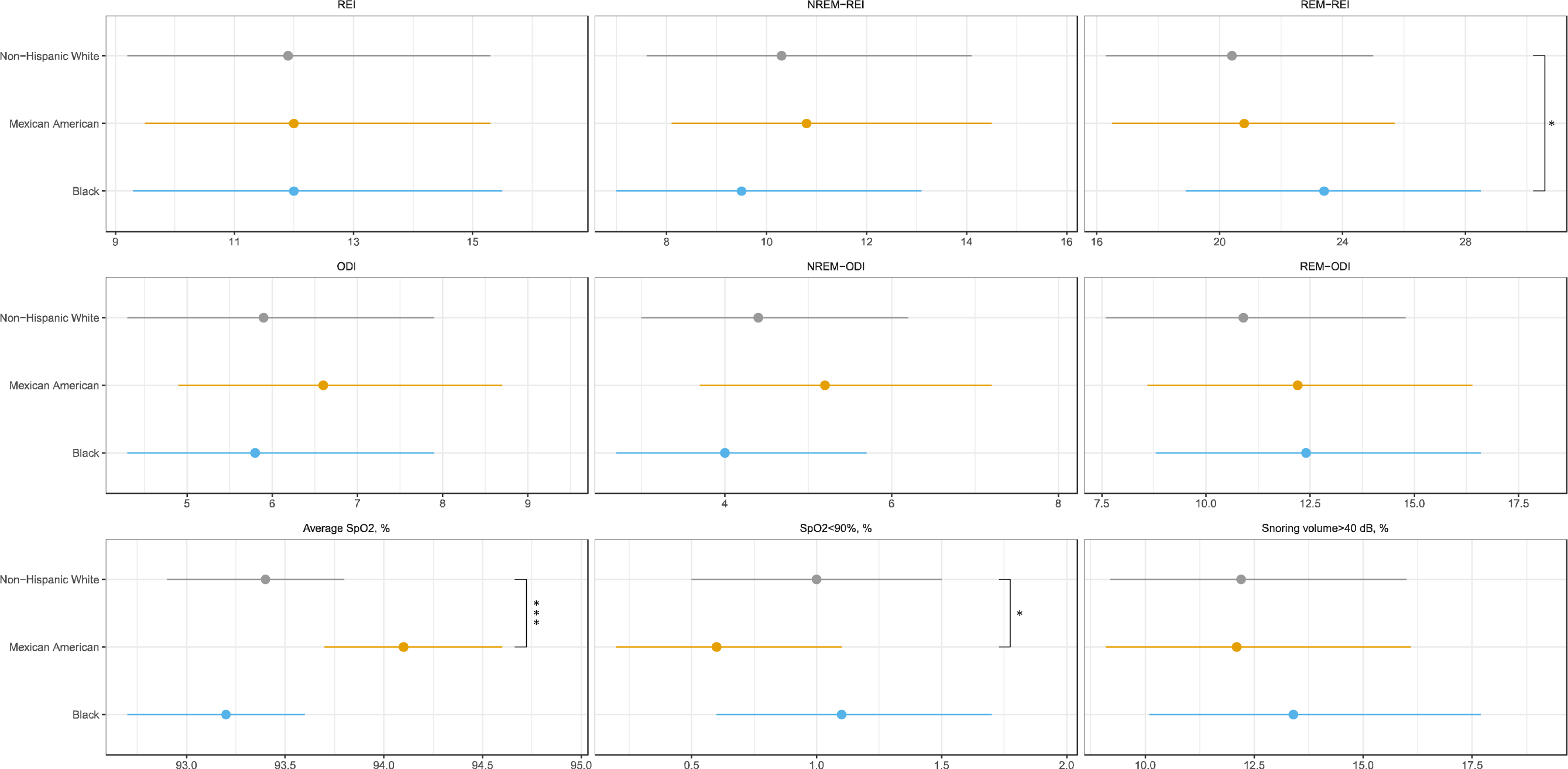
Multivariable-adjusted mean sleep-disordered breathing metrics by race. REI: Respiratory Event Index; REM: Rapid eye movement; NREM: Non-rapid eye movement; ODI: Oxygen desaturation index Models were adjusted for age, sex, education, income, employment status, cognitive status, body mass index, history of hypertension, diabetes, stroke, and coronary heart disease, and sleep medication use. Comparisons were made using the Non-Hispanic White individuals as the reference group. *<0.05, ** <0.01, ***<0.001.

## DISCUSSION

In this community-based cohort of 821 middle- to older-aged NHW, MA, and Black adults, objective in-home measures indicate a high prevalence of moderate to severe SDB, especially during REM sleep. While the overall prevalence of moderate to severe SDB (including all events across the entire sleep period) did not differ by race or sex, we observed distinct patterns of REM and NREM SDB measures among different subgroups of the population. Black women had the highest REM REI and the highest proportion of snoring volume>40 dB; MA women had the highest average SpO2 levels; and NHW men had the lowest REM REI and REM ODI. Compared to NHW participants, Black participants had significantly higher REM REI, and MA participants had higher blood oxygen levels, independent of socioeconomic factors, BMI and several comorbidities.

Few studies have examined SDB prevalence in diverse populations. This is clinically important, particularly given the low rates of diagnosis and treatment of SDB in resource-poor settings, which may impose a significant health burden on minority populations^4^. The Jackson Heart Health Study showed an obstructive sleep apnea (OSA) prevalence of 53.6% among African American adults aged 50 to 80 years, and a prevalence of 20.4% for moderate to severe OSA^8^. In a cohort of diverse participants aged 54-93, a significantly higher prevalence was reported for OSA among Hispanic adults compared to NHW individuals, but not among Black adults^6^. In the current study, 87.8% of the sample had SDB, and about half had moderate to severe SDB, including 72.7% moderate to severe REM SDB, as measured by a PAT-based home testing device. While the high rates observed in this study could relate to over-estimation of SDB using the WatchPAT^22^, we only included SDB events that were manually annotated and met clear desaturation criteria. The high prevalence of moderate to severe SDB, particularly REM SDB, may also stem from the unfavorable metabolic profiles^23^ of this study population, with a median BMI of 29.8, and 63.9% of the sample had a history of hypertension and 25.5% had a history of diabetes, respectively. Over 20% of our participants had MCI or dementia, which may also result in a higher prevalence of SDB, given the close link between SDB and cognitive aging^24,25^.

Despite significantly higher rates of hypertension, diabetes, and cognitive impairment among Black and MA adults compared to NHW adults, the overall REIs (combining REM and NREM) were similar across the three race groups in our study. It has been suggested that racial differences in SDB severity may arise from different contributions of various SDB mechanistic traits to propensity for airway collapse, which also vary by age^26^. Specifically, these racial differences may be more evident in individuals under 25, with SDB severity decreasing in older Black adults, leading to similar SDB severity between middle-aged and older Black and NHW adults, potentially due to greater contributions of anatomic features (soft tissue, adenotonsillary hypertrophy) to SDB in younger individuals ^26–28^.

Importantly, however, our study showed for the first time that community-dwelling Black-but not MA participants-had significantly higher REM REI compared to NHW adults, independent of socioeconomic factors, BMI, and several comorbidities. While our findings align with the only other study reporting racial and ethnic differences in REM OSA-showing a higher prevalence of REM-related OSA among African American patients than non-African Americans^29^-that previous study was conducted in a clinical sample of patients referred to a sleep laboratory for suspected OSA, limiting its generalizability to broader populations. Differences in the endotypic characteristics may explain the similar overall REIs but differing REM REIs observed between Black and NHW adults. For instance, decreased airway collapsibility in Black individuals may reflect relative protection against respiratory events in NREM sleep, leading to similar overall AHIs, particularly in older Black and White adults ^27^. It is also postulated that hypopneas may be under-recognized in darker pigmented individuals due to biases in oximeters^30^. During REM sleep, where oxygen desaturations are typically deeper, hypopneas may be more easily detected, possibly facilitating identification of events in REM sleep despite under-estimation of desaturation due to oximetry bias. Additionally, Black participants in our sample were notably younger and more likely to be women, which are both strong correlates of REM OSA^9,31–33^.

Since BMI has considerably greater effects on oxygen desaturation during REM sleep, it is plausible that the significantly higher prevalence of obesity among Black participants in our sample contributed to greater degree of nocturnal hypoxemia and higher REM REI. Notably, the racial differences in REM REI remained robust after accounting for socioeconomic factors, BMI, cognitive diagnoses, a history of hypertension, diabetes, stroke, and CHD, sleep medication use, and sleep position. It has been suggested that patients with REM SDB may be more symptomatic, often with longer events, greater hypoxemia, and higher levels of sympathetic activity, compared to NREM SDB, despite having an average REI (or AHI) falling within the normal range^34,35^. Consequently, REM SDB has been associated with higher cardiovascular risk, including an increased risk of hypertension, metabolic syndrome, and diabetes^31,36^, and recurrent cardiovascular events in those with prevalent cardiovascular diseases^16^. Our findings emphasize the importance of assessing SDB, particularly REM SDB, in minoritized populations.

Interestingly, among the six sex-race combined groups, while men and women did not differ in SDB characteristics overall, Black women had the highest REM REI and the highest proportion of snoring volume>40 dB, and NHW men had the lowest REM REI and REM ODI. It has been reported that the prevalence of OSA is approximately twice as common in men as in women, although these sex differences in OSA tend to decrease with age, as the prevalence has been reported to double in women after menopause^37,38^. Additionally, women have been found to have significantly higher prevalence of REM-predominant OSA than men^39^. This may be due to the reduced protective effects of female sex hormones on upper airway muscles during REM sleep, when atonia occurs, or due to the smaller size of the pharyngeal wall in women, making them more susceptible to airway collapse during REM sleep^32,39^. OSA has long been considered a male-predominant disease and has been understudied in women^40^. Our divergent findings on REM SDB measures between NHW and Black women emphasize the critical needs to consider both sex and race, as well as stage-specific SDB measures, to gain a more nuanced understanding of the disease. Further research is needed to clarify the mechanisms that may explain state-specific differences in SDB across race, ethnic, and sex groups, considering social determinants of health and factors associated with gender that may influence SDB pathophysiology.

This study has several strengths. The use of a low-burdensome HSAT device to characterize nighttime sleep quality and OSA at home in a community-based diverse cohort provides a unique opportunity to clarify sleep health disparities in real-world settings. In particular, this is among the first study, to our knowledge, to examine differences in sleep state-specific OSA measures among Black, MA, and NHW adults, taking into account of the influence of socioeconomic factors as well as multiple comorbidities. This is critical for elucidating the clinical implications of REM and NREM SDB, including their impact on health in diverse populations.

There are also a few limitations. While WatchPAT is an FDA-approved home sleep apnea test device, it has not been specifically validated in race/ethnic minority groups, including Black or MA participants. There have been suggestions that the accuracy of photoplethysmography (PPG), a method commonly used in sleep wearable devices to evaluate heart rate and peripheral blood oxygen saturation based on red and infrared light absorption levels, may overestimate oxygen levels among populations with dark skin tones^30,41^. This may have contributed to the higher average SpO2 levels observed among MA participants, although this effect was not noted among Black individuals. Sleep was not assessed using neurophysiological assessment and it is likely that there is some misclassification due to indirect measurement of REM sleep. However, we are unaware of factors that would result in non-differential misclassification, and thus, findings may be biased to the null. Nevertheless, further research is required to examine the accuracy of WatchPAT for assessing sleep staging and SDB metrics in Black and MA adults. Additionally, participants were only assessed for one night, and thus we were unable to examine night-to-night variability in SDB in this diverse population.

## Conclusion

In this community-based cohort of middle- to older-aged NHW, MA, and Black adults, PAT-based measures of in-home sleep indicate a higher prevalence of REM SDB in Black individuals, particularly Black women, independent of age, sex, education, income, employment status, cognitive status, BMI, and several comorbidities. Home-based measures that do not distinguish between REM and NREM respiratory events may underestimate sleep apnea in racial minority groups. Given the close link between REM SDB and elevated cardiovascular disease risk, clinicians should pay more attention to this sleep apnea phenotype, especially in minoritized populations.

## Data Availability

Data is available upon request from the authors.

## Acknowledgements

YL had full access to all of the data in the study and takes responsibility for the integrity of the data and the accuracy of the data analysis. YL, CC, CP, SEO, SR, and KY contributed substantially to the study design, data analysis and interpretation, and the writing of the manuscript.

## Abbreviations

BMI: body mass index
CHD: coronary heart disease
HABS-HD: Health and Aging Brain Study-Health Disparities
MA: Mexican American
MCI: mild cognitive impairment
NHW: non-Hispanic White
NREM: non-rapid eye movement
ODI: oxygen desaturation index
OSA: obstructive sleep apnea
PAT: Peripheral Arterial Tonometry
REM: rapid eye movement
REI: respiratory event index
SDB: sleep-disordered breathing
SES: socio-economic status
CBPR: community-based participatory research

