## Supplemental tables for "Racial and ethnic and sex differences in at-home estimates of sleep-disordered breathing parameters among Mexican American, Black, and Non-Hispanic White adults"

Supplemental table 1. Description of participants characteristics by race and ethnicity

|  | Total sample  (n=821) | Non-Hispanic White  (n=363) | Mexican American  (n=284) | Black  (n=174) | p-value^a^ |
| --- | --- | --- | --- | --- | --- |
| Female | 543 (66.1) | 217 (59.8) | 203 (71.5) | 123 (70.7) | 0.003 |
| Age, years* | 66.6 (±8.5) | 69.9 (±8.0) | 64.4 (±7.0) | 63.2 (±7.8) | <.0001 |
| MCI/dementia | 176 (21.4) | 52 (14.3) | 74 (26.1) | 50 (28.7) | <.0001 |
| BMI, kg/m^2^ | 29.8 (26.1,34.6) | 29.1 (24.6,33.0) | 29.8 (26.8,35.1) | 32.1 (27.4,37.4) | <.0001 |
| Obesity (BMI≥30 kg/m^2^ ) | 397 (48.9) | 156 (43.2) | 136 (48.6) | 105 (61.4) | 0.0005 |
| History of hypertension | 525 (63.9) | 211 (58.1) | 175 (61.6) | 139 (79.9) | <.0001 |
| History of diabetes | 209 (25.5) | 55 (15.2) | 101 (35.6) | 53 (30.5) | <.0001 |
| History of stroke | 40 (4.9) | 23 (6.3) | 7 (2.5) | 10 (5.7) | 0.06 |
| History of heart attack | 31 (3.8) | 13 (3.6) | 9 (3.2) | 9 (5.2) | 0.53 |

^*^ mean (± standard deviation); all other results are shown as n (%) or median (Q1,Q3)

MCI: Mild cognitive impairment; BMI: Body mass index ^a^ Mann-Whitney U test was used for continuous variables, Chi-square test for categorical variables

Supplemental table 2. Description of sleep-disordered breathing parameters by sex

| **Sleep-disordered breathing variables** | Total sample  (n=821) | Male  (n=278) | Female  (n=543) | p-value^a^ |
| --- | --- | --- | --- | --- |
| REI | 15.0 (8.5,25.1) | 15.0 (7.4,27.7) | 15.1 (9.0,23.9) | 0.99 |
| NREM-REI | 11.7 (5.9,20.8) | 13.0 (5.6,25.7) | 11.1 (6.0,19.8) | 0.10 |
| REM-REI | 25.1 (13.6,37.45) | 24.1 (10.7,37.9) | 25.8 (15.0,37.4) | 0.08 |
| ODI | 7.1 (3.1,14.5) | 7.2 (2.7,17.4) | 7.1 (3.3,13.5) | 0.60 |
| NREM ODI | 4.6 (1.9,10.6) | 4.7 (1.8,14.0) | 4.6 (2.0,9.5) | 0.36 |
| REM-ODI | 13.3 (5.7,25.85) | 12.3 (4.9,25.9) | 14.0 (6.4,25.8) | 0.15 |
| Average SpO2, %* | 93.7 (±1.6) | 93.6 (±1.6) | 93.7 (±1.7) | 0.32 |
| SpO2<90%, % | 0.3 (0.0,1.6) | 0.3 (0.0,2.1) | 0.2 (0.0,1.5) | 0.53 |
| Snoring volume>40 dB, % | 13.4 (6.1,28.3) | 14.1 (7.3,25.9) | 13.0 (5.6,29.6) | 0.54 |

^*^ mean (± standard deviation). All other variables are presented as median (Q1,Q3).

REI: Respiratory Event Index; REM : Rapid eye movement; NREM: Non-rapid eye movement; ODI: Oxygen desaturation index

^a^ Kruskal-Wallis test was used for continuous variables with a normal distribution and Student’s t-test for continuous variables without a normal distribution, Chi-square test for categorical variables

Supplemental table 3. Description of sleep-disordered breathing parameters by sex and race and ethnicity

| **Sleep-disordered breathing variables** | **Men (n=278)** | | | | **Women (n=543)** | | | |
| --- | --- | --- | --- | --- | --- | --- | --- | --- |
|  | NHW (n=146) | MA (n=81) | Black (n=51) | p^a^ | NHW (n=217) | MA (n=203) | Black (n=123) | p^a^ |
| REI | 14.7 (6.7,26.8) | 17.0 (7.6,29.0) | 17.0 (8.1,27.2) | 0.38 | 14.3 (8.4,23.8) | 14.9 (9.1,24.7) | 16.0 (10.6,23.7) | 0.41 |
| NREM-REI | 11.7 (5.4,25.1) | 14.7 (6.4,27.5) | 13.9 (5.9,23.6) | 0.48 | 10.5 (6.0,20.0) | 11.2 (5.8,19.7) | 11.3 (6.6,18.3) | 0.97 |
| REM-REI | 21.7 (9.0,34.0) | 27.8 (11.7,38.2) | 26.2 (15.1,45.9) | 0.02 | 24.2 (13.7,36.1) | 26.4 (15.7,38.5) | 28.0 (18.4,38.8) | 0.04 |
| ODI | 6.3 (2.3,15.1) | 7.9 (3.5,19.7) | 5.6 (2.7,15.4) | 0.14 | 6.2 (3.1,12.8) | 8.5 (3.8,14.7) | 7.3 (3.9,12.8) | 0.22 |
| NREM-ODI | 4.3 (1.5,10.9) | 5.8 (2.7,19.0) | 3.8 (2.1,11.6) | 0.12 | 4.1 (1.9,10.0) | 5.3 (2.2,10.2) | 4.8 (1.9,8.7) | 0.46 |
| REM-ODI | 9.5 (3.3,21.1) | 15.8 (6.6,27.5) | 15.1 (6.1,28.1) | 0.03 | 12.6 (5.2,23.8) | 15.9 (6.5,28.9) | 14.2 (7.6,24.9) | 0.11 |
| Average SpO2, %* | 93.6 (±1.4) | 94.0 (±1.9) | 93.2 (±1.5) | 0.02 | 93.5 (±1.7) | 94.3 (±1.4) | 93.3 (±1.6) | <.0001 |
| SpO2<90%, % | 0.3 (0.0,1.5) | 0.5 (0.1,2.5) | 0.4 (0.0,4.2) | 0.11 | 0.3 (0.0,1.6) | 0.2 (0.0,1.2) | 0.3 (0.1,1.8) | 0.049 |
| Snoring volume>40 dB, % | 12.1 (6.8,24.5) | 15.6 (7.7,27.7) | 14.9 (7.8,33.4) | 0.23 | 10.4 (5.0,23.8) | 14.7 (5.7,33.2) | 18.2 (7.2,35.3) | 0.003 |

^*^ mean (± standard deviation). All other variables are presented as median (Q1,Q3).

REI: Respiratory Event Index; REM : Rapid eye movement; NREM: Non-rapid eye movement; ODI: Oxygen desaturation index; NHW: Non-hispanic White; MA: Mexican American

^a^ Kruskal-Wallis test was used for continuous variables with a normal distribution and Student’s t-test for continuous variables without a normal distribution, Chi-square test for categorical variables

Supplemental table 4. Multivariable-adjusted sleep-disordered breathing parameters by race and ethnicity

| **Sleep-disordered breathing variables** | Non-Hispanic White  (n=363) | Mexican American  (n=284) | Black  (n=174) | p-value | Direction |
| --- | --- | --- | --- | --- | --- |
| REI | 11.9 (9.2,15.3) | 12.0 (9.5,15.3) | 12.0 (9.3,15.5) | 0.99 | - |
| NREM-REI | 10.3 (7.6,14.1) | 10.8 (8.1,14.5) | 9.5 (7.0,13.1) | 0.51 | - |
| REM-REI | 20.4 (16.3,25.0) | 20.8 (16.5,25.7) | 23.4 (18.9,28.5) | 0.14 | NHW< B |
| ODI | 5.9 (4.3,7.9) | 6.6 (4.9,8.7) | 5.8 (4.3,7.9) | 0.42 | - |
| NREM-ODI | 4.4 (3.0,6.2) | 5.2 (3.7,7.2) | 4.0 (2.7,5.7) | 0.10 | MA >B |
| REM-ODI | 10.9 (7.6,14.8) | 12.2 (8.6,16.4) | 12.4 (8.8,16.6) | 0.42 | - |
| Average SpO2, % | 93.4 (92.9,93.8) | 94.1 (93.7,94.6) | 93.2 (92.7,93.6) | <.0001 | MA> NHW,B |
| SpO2<90%, % | 1.0 (0.5,1.5) | 0.6 (0.2,1.1) | 1.1 (0.6,1.7) | 0.04 | MA< NHW,B |
| Snoring volume>40 dB, % | 12.2 (9.2,16.0) | 12.1 (9.1,16.1) | 13.4 (10.1,17.7) | 0.57 | - |

All variables are presented as adjusted means (95%CI).

The model adjusted for age, sex, education, employment status, cognitive status, physical activity, smoking, body mass index, history of hypertension, diabetes, stroke, and heart attack, and sleep medication use.

REI: Respiratory Event Index; REM : Rapid eye movement; NREM: Non-rapid eye movement; ODI: Oxygen desaturation index; NHW: Non-hispanic White; MA: Mexican American.

Variables lack of gaussian distribution were applied a transformation and then back-transformed to the original scale to show the results.
